# Real-time SARS-CoV-2 diagnostic and variants tracking over multiple candidates using nanopore DNA sequencing

**DOI:** 10.1101/2021.05.18.21257281

**Authors:** François Stüder, Jean-Louis Petit, Stefan Engelen, Marco Antonio Mendoza-Parra

## Abstract

Since December 2019, the emergence of a novel coronavirus responsible for a severe acute respiratory syndrome (SARS-CoV-2) is accountable for a major pandemic situation. As consequence, a major effort worldwide has been performed for the development of viral diagnostics strategies aiming at (i) reducing the diagnostics time, (ii) decrease the costs per assay, and (iii) providing population-scale solutions.

Beyond the diagnostics requirements, the description of the B.1.1.7 strain, originated in the south of England, as a highly transmissible variant has strongly accelerated the world-wide interest in tracking SARS-CoV-2 variants’ occurrence. Since then, other extremely infectious variants, were described and unsurprisingly further others are expected to be discovered, notably due to the long period of time on which the pandemic situation is lasting.

Interestingly, all currently described SARS-CoV-2 variants present systematically several mutations within the gene encoding the Spike protein, involved in host receptor recognition and entry into the cell. Hence, instead of sequencing the whole viral genome for variants’ tracking, our proposed strategy focuses on the SPIKE region, as a way to increase the number of candidate samples to screen at once; an essential aspect to accelerate SARS-CoV-2 diagnostics, but also improve variants’ emergence and progression surveillance.

Herein we present a proof of concept study, for performing both at once, population-scale SARS-CoV-2 diagnostics and variants’ tracking. This strategy relies on (i) the use of the portable and affordable MinION DNA sequencer; (ii) a DNA barcoding strategy and a SPIKE gene-centered variant’s tracking, for largely increasing the number of candidates per assay; and (iii) a real-time diagnostics and variant’s tracking monitoring thanks to our software RETIVAD.

As a whole, this strategy represents an optimal solution for addressing the current needs on SARS-CoV-2 progression surveillance, notably due to its affordable implementation, allowing its implantation even in remote places over the world.

## Introduction

Since December 2019, the emergence of a novel coronavirus responsible for a severe acute respiratory syndrome (SARS-CoV-2) is accountable for a major pandemic situation, leading to date to nearly 3 million deaths worldwide. Early on 2020 major viral diagnostic efforts were deployed, including the development of news strategies aiming at (i) reducing the diagnostics time, (ii) decrease the costs per assay, (iii) providing population-scale solutions, but also (iv) presenting a high sensitivity and specificity, notably to discriminate asymptomatic cases. While costs and required time per assay were early on addressed by the development of immune-based tests ^1^, their sensitivity relative to viral nucleic acid-targeting diagnostics were initially questioned, but finally accepted in several countries notably due to the low throughput of diagnostics based on RT-qPCR assays ^2^.

With the aim of taking advantage of the viral nucleic acid targeting diagnostics but at the same time increasing the number of candidates’ through-put, strategies based on the use of massive parallel DNA sequencing for population-scale diagnostics were proposed ^3–5^. These efforts rely on (i) the incorporation of DNA molecular barcodes during sample preparation; (ii) pooling barcoded samples for an “in-one-run” diagnostics assay by the use of massive parallel DNA sequencing; (iii) stratification of diagnostics’ outcome with the help of bioinformatics processing. While such strategies demonstrated to up-scale the diagnostics power beyond several thousand candidates per assay, they require to count with a DNA sequencing platform where large instruments (Illumina Sequencers) are driven by specialized technical engineers. As consequence, this strategy remains applicable to highly developed countries (due to the prohibitive costs of the required DNA sequencers), which in addition requires a dedicated pipeline strategy for transporting collected samples towards sequencing centers.

Beyond the diagnostics requirements, the identification of the highly transmissible SARS-CoV-2 variant B.1.1.7 - originated in the south of England - has strongly accelerated the worldwide interest in tracking SARS-CoV-2 variants’ occurrence. More recently, two other extremely infectious variants, namely the P.1 issued on the Brazilian city of Manaus and the B.1.351 initially detected In South Africa, were described, and unsurprisingly further others are expected to be discovered, notably due to the long period of time on which the pandemic situation is lasting ^6^.

Herein, we describe a proof of concept study for population-scale diagnostics and variants’ tracking using massive parallel DNA sequencing performed with the MinION Oxford Nanopore Technology (ONT). ONT MinION sequencers are well-known for their portability, their reduced cost, and their simplicity of use; thus, representing a suitable strategy for its implementation even in remote places over the world. In addition to a dedicated molecular biology protocol, we deliver a computational solution (RETIVAD: Real-Time Variants Detector) providing the outcome of the diagnostic in a real-time mode during sequencing; allowing to earn time for diagnostics report, essential in the current context of the pandemic situation. Finally, we have also demonstrated that our strategy is able to perform SARS-CoV-2 variants’ detection across multiple candidates, notably by targeting the SPIKE gene instead of covering the whole viral genome (**Figure 1**). This targeted variants’ tracking strategy allows increasing ∼10x the potential sequencing coverage delivered during the assay, thus allowing to redistribute this resource towards the variants’ screening of a large number of candidates at once.

**Figure 1.**
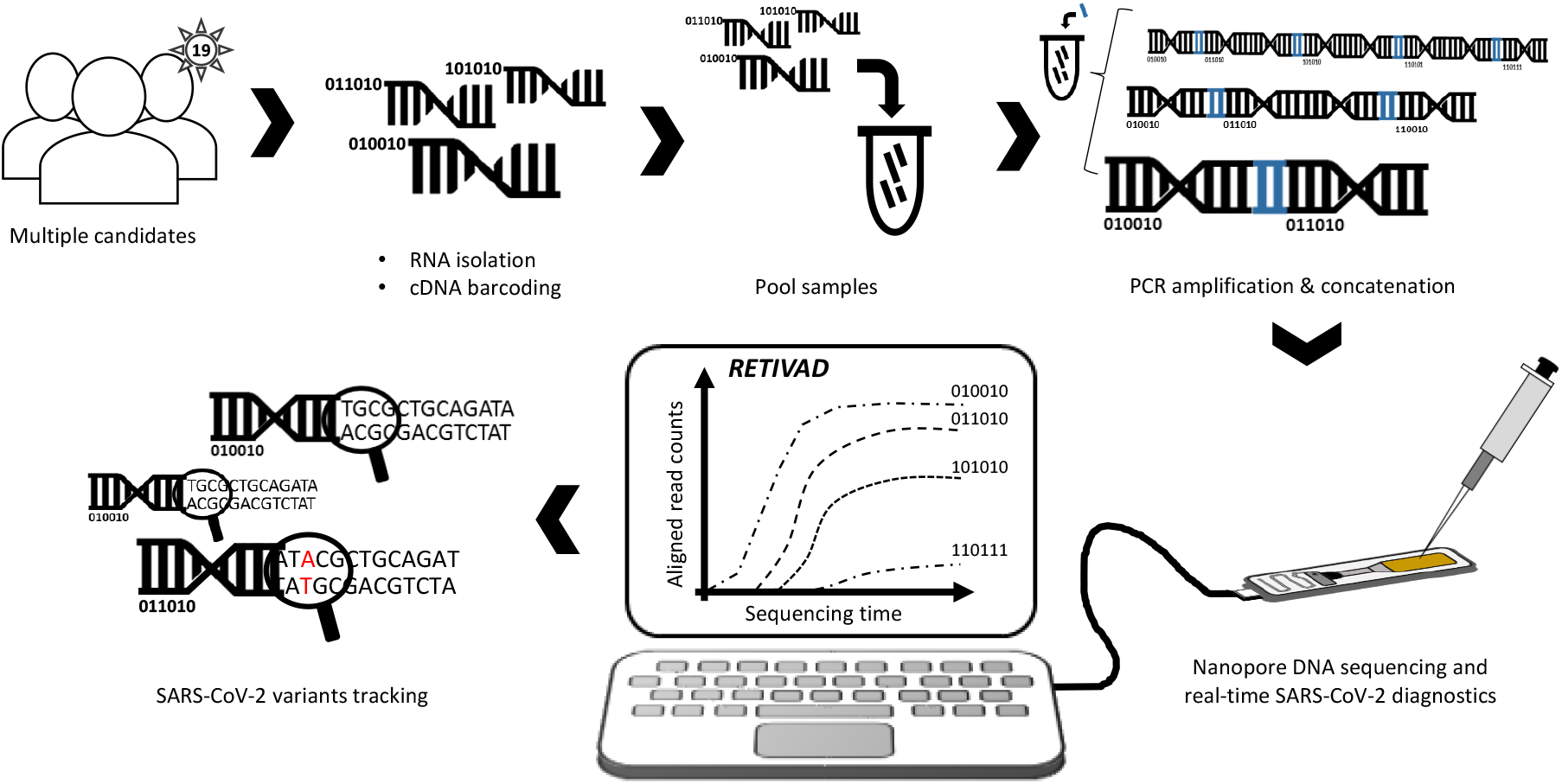
Scheme illustrating the proposed real-time SARS-CoV-2 diagnostics and variants tracking strategy. RNA samples isolated from multiple candidates are converted into complementary DNA (cDNA) such that samples associated to each candidate are labelled by molecular DNA barcodes appended to their extremities. Barcoded-cDNA samples are pooled together and PCR amplified in presence of a bridge sequence (blue) allowing to generate concatemers. Long DNA concatemers are loaded into the MinION nanopore sequencer for real-time SARS-CoV-2 diagnostics and variants tracking with the help of RETIVAD.

In summary, the proposed strategy satisfies the current need for counting with a population diagnostic assay, and discriminates between the various SARS-CoV-2 variants, which are recognized to potentially present variable infection properties as well as clinical outcomes ^6,7^.

## Results

### A combinatorial molecular barcode strategy for SARS-CoV-2 diagnostics over multiple candidates

With the aim of performing SARS-CoV-2 diagnostics over multiple candidates with the MinION sequencer, we combined a regular reverse-transcription assay, with a particular PCR amplification allowing the generation of long DNA concatemers. This strategy allows to target short amplicons, like those used on quantitative real-time PCR strategies, but at the same time take advantage of the long fragments sequencing capabilities of the Oxford Nanopore technology.

Reverse transcription (RT) is performed with a Covid-19 specific primer, presenting, in addition, a unique molecular barcode (20nt length) and a common sequence adapter (Gibson sequence; 30nt length) (**Figure 2A**). Hence, multiple candidate’ samples are reverse-transcribed with a unique molecular barcode per candidate, allowing their identification after massive parallel DNA sequencing.

**Figure 2.**
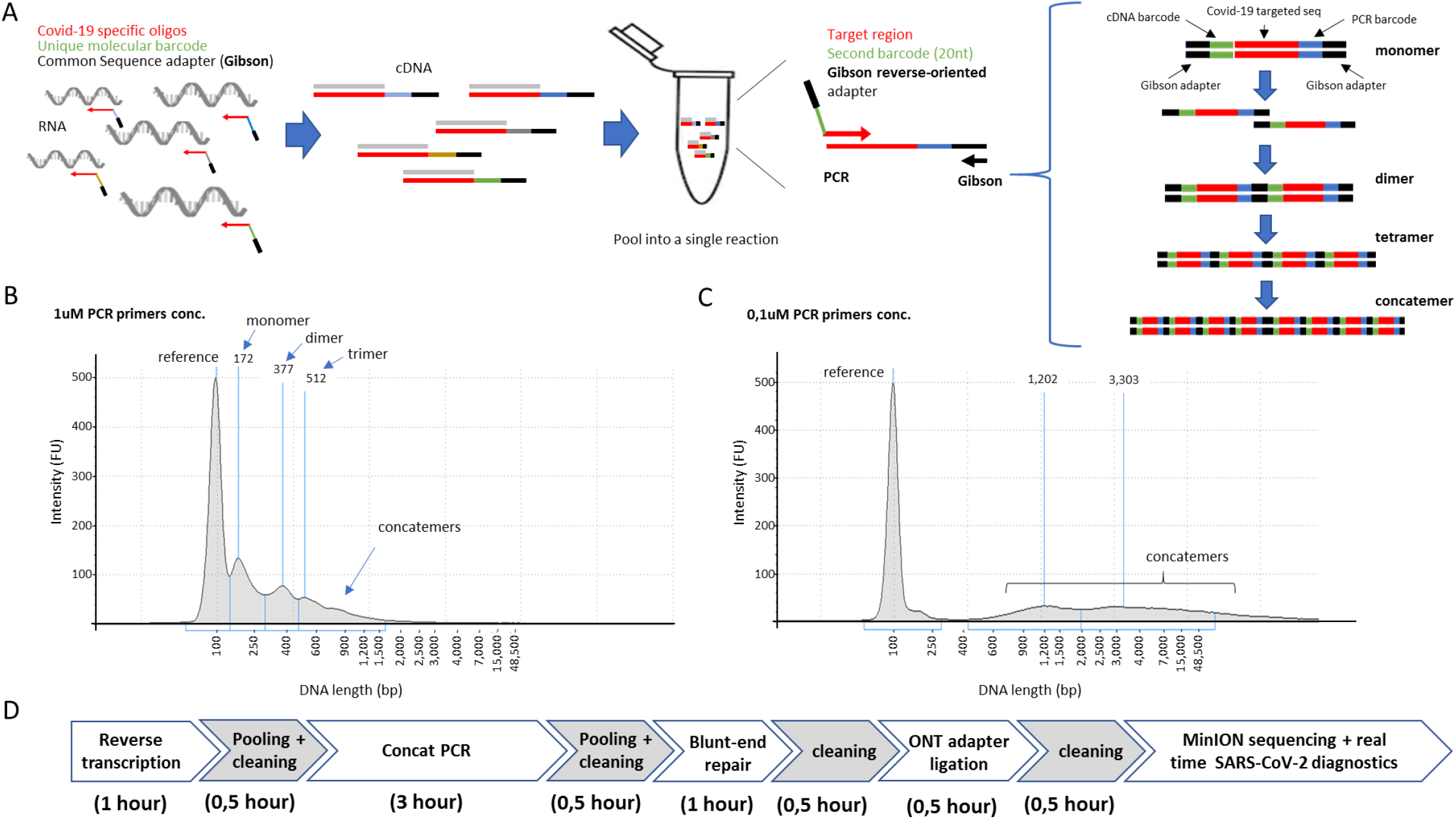
Dual molecular DNA barcoding and PCR-driven concatemerization applied to SARS-CoV-2 diagnostics. **(A)** Reverse transcription is performed with oligonucleotides targeting a Covid-19 region, but in addition containing a unique molecular barcode per sample and a common adapter sequence (Gibson). Reverse transcription assays (cDNA) performed for multiple candidates are pooled together and PCR amplified by using an oligonucleotide targeting the common Gibson sequence as well as a second Covid-19-specific primer hosting a second molecular barcode and a complementary Gibson sequence. During PCR cycles, amplified products are able to concatenate thanks to the Gibson flanking sequences, hence generating concatemers. We have called this particular PCR strategy “Concat-PCR”. **(B)** DNA electropherogram (Agilent TapeStation system) illustrating the various PCR concatemer products obtained with PCR primers at 1uM final concentration. **(C)** DNA electropherogram for a concat-PCR assay performed with PCR primers at 0.1uM final concentration, demonstrating a preferential production of long concatemers in contrast to those observed in (B). (D) Timeline illustrating the various molecular biology steps required for samples preparation prior engaging samples into MinION DNA sequencing.

After RT assay, candidates’ samples are pooled for PCR amplification into a single reaction. For it, a second Covid-19 specific primer, presenting, in addition, a unique molecular barcode (20nt length) and a reverse-oriented sequence for the common Gibson sequence adapter (30nt length) is used together with a Gibson primer (**Figure 2A**). This strategy allows to incorporate a second unique molecular barcode, thus providing a combinatorial barcoding strategy per candidate which is extremely useful for increasing the number of candidates to interrogate without having to linearly increase the number of primers to purchase; an aspect which is of major relevance due to their long length (>70nt).

The presence of Gibson sequences on both extremities of the PCR amplified products allows generating concatemers during amplification (**Figure 2A**). While this process takes place in an uncontrolled manner during PCR amplification, low primer concentrations were shown to give rise preferentially to long concatemers (**Figure 2B & 2C**). As consequence, concatenated PCR products can be purified from long primers (>70nt length) with SPRI-select reagent (Beckman; B23318), without the risk of losing the short amplicons. Furthermore, the use of long concatemers increased the average sequencing coverage of MinION by a factor of ∼9 (i.e. 1 million concatemers sequenced corresponds to 9 million monomers; **Figure S1 and Figure 3E**), such that a large number of candidates can be potentially screened like in previous population-scale diagnostics efforts based on the use of Illumina instruments ^3–5^.

**Figure 3.**
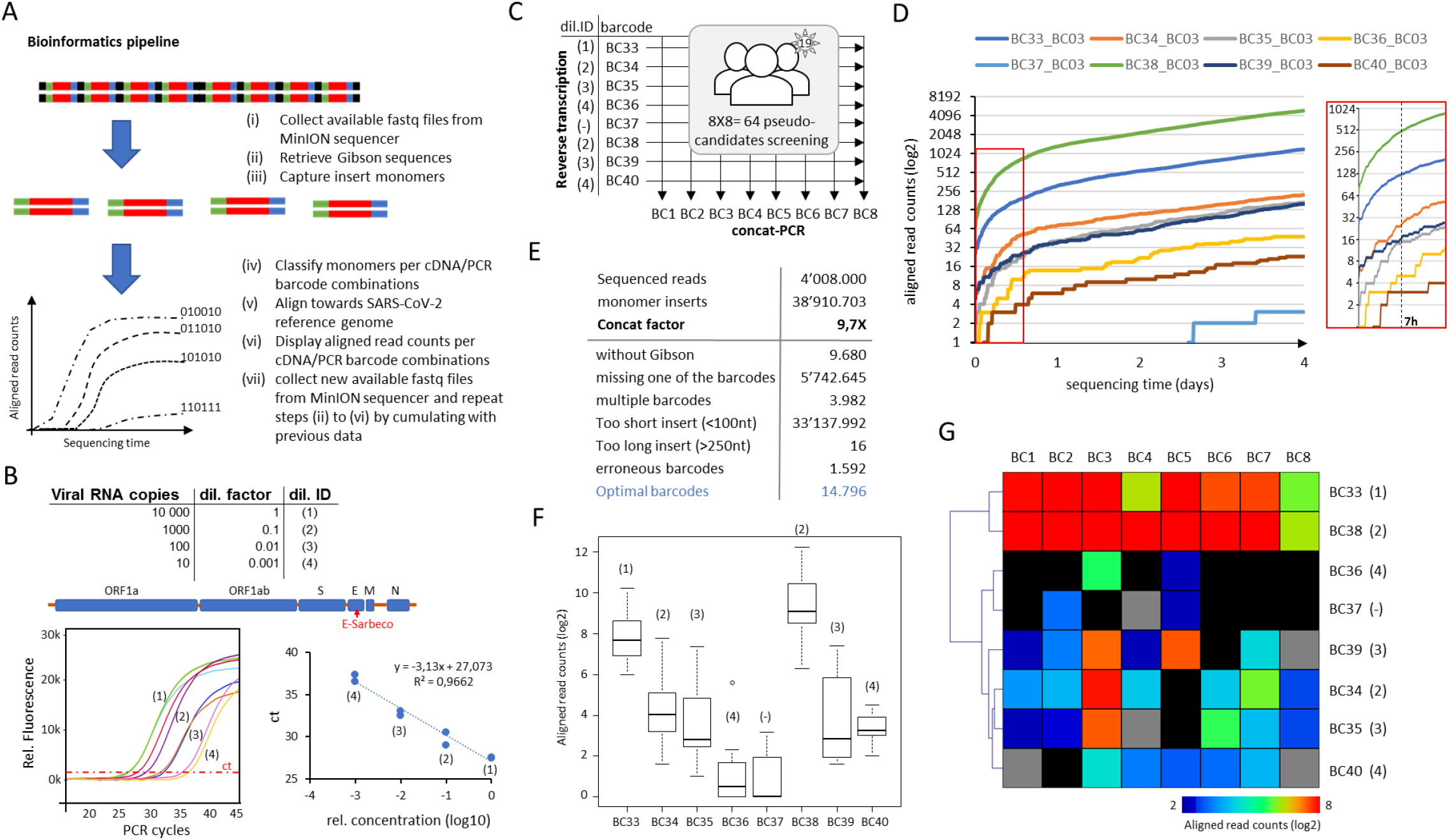
SARS-CoV-2 diagnostics assay with Nanopore DNA sequencing. **(A)** Scheme of the bioinformatics pipeline within RETIVAD. Long concatemer sequenced reads are computationally fragmented in monomers with the help of the Gibson sequence location. Monomers are stratified on the basis of flanking barcodes (cDNA and PCR incorporated barcodes) and aligned to the Covid-19 viral reference genome to verify its specificity. Considering that Oxford Nanopore instruments provide sequenced data during sequencing, the aforementioned steps are performed on real-time, such that the number of aligned read counts per candidate (defined by the combination of cDNA & PCR incorporated DNA barcodes) are assessed and displayed during the MinION sequencing process. **(B)** The synthetic SARS-CoV-2 RNA control sample (Twist Bioscience; Wuhan-Hu-1 strain) was subsequently diluted and used for reverse transcription and RT-qPCR assay targeting the E-Sarbeco viral region. **(C)** Reverse transcription assay scheme for 8 conditions corresponding to different viral RNA dilutions (dilution ID corresponding to that presented in (B)) and targeting the E-Sarbeco region. Each of these reverse transcription assays are identified by the indicated molecular barcodes (BC33-BC40). Note that among them a sample devoid of viral RNA is also included (BC37). All 8 samples are pooled at the end of the assay and concat-PCR amplified with a set of other eight barcoded primers targeting the E-Sarbeco gene (BC1-BC8), hence providing a combinatorial barcoding strategy corresponding to 64 pseudo-candidates. **(D)** Real-time display illustrating the number of aligned read counts associated to the E-Sarbeco region per dual barcode combination (BC3 vs all cDNA barcodes) assessed during Nanopore sequencing. Inset: Zoom view highlighting that the first seven hours of DNA sequencing are sufficient to reveal read count differences between samples. **(E)** Summary of the bioinformatics pipeline assessed at the end of 4 days of DNA sequencing. Concatenation factor is inferred from the ratio between the number of detected monomers and the initial number of sequenced concatemers. Optimal barcodes correspond to the number of sequences presenting an optimal monomer length (>100nt; <250nt) and presenting both cDNA & PCR barcode sequences. **(F)** Boxplots illustrating the total aligned read counts associated to each cDNA barcode (BC33-BC40) and the different PCR barcodes. Each of the boxplots are labelled with their corresponding viral RNA dilution (dil.ID indicated in (B)). **(G)** Heatmap illustrating the total aligned read counts associated to each cDNA barcode (BC33-BC40) and the different PCR barcodes (BC1-BC8). Note that (F) & (G) reveals a sensitivity of 100 viral copies, since the next RNA dilution condition appears in some cases at the same level of the negative samples.

Overall, the proposed pipeline is composed of a standard molecular biology strategy, requiring less than 8 hours of samples preparation prior Nanopore sequencing (**Figure 2D** and Methods section).

### Real-time SARS-CoV-2 diagnostics over multiple pseudo-candidates presenting variable amounts of viral copies

One of the major bottlenecks for population-scale Covid-19 diagnostics based on massive parallel DNA sequencing is the required time to deliver the outcome of the diagnostic. In addition to the molecular biology workflow required for samples’ processing till sequencing library preparation, DNA sequencing, performed on Illumina instruments, requires an incompressible processing time, which delays downstream analyses since they can only be performed at the end of the whole sequencing process. In contrary, Oxford Nanopore sequencers deliver sequenced reads on a rolling basis (4000 sequenced reads per generated fastq file). Hence, we have developed a computational pipeline dedicated to (i) collect available fastq files; (ii) retrieve Gibson adapter sequences within sequenced molecules; (iii) split them into monomers; (iv) evaluate the presence of cDNA & PCR molecular barcodes in their flanking regions; (v) align the inner monomer sequences towards the SARS-CoV-2 reference genome; and (vi) display the aligned read counts per cDNA/PCR barcode combinations. This pipeline is setup to be reinitiated in an automated time interval (e.g. every 10 minutes) such that newly available sequences can be cumulated to the previous analyses, thus providing a real-time view of aligned read counts associated with cDNA/PCR barcode combinations (**Figure 3A**).

To validate the performance of the proposed diagnostics strategy, synthetic SARS-CoV-2 RNA control samples (Twist Biosciences) were used for mimicking variable viral RNA copies per assay. Specifically, subsequent dilutions of the synthetic Wuhan-Hu-1 (GeneBankID: MN908947.3) SARS-CoV-2 RNA control sample has been used for reverse transcription, followed by a quantitative PCR assay targeting a region on the E gene of SARS-Cov-2, defined by primer sequences described previously (E-Sarbeco) ^8^. This quantitative PCR allowed to detect as few as 10 viral copies of the synthetic SARS-CoV-2 RNA control sample (**Figure 3B**).

For Nanopore DNA sequencing-based diagnostic, as described on **Figure 2A**, we have designed eight barcoded E-Sarbeco reverse primers (targeting the same amplicon region used for the quantitative PCR assay), and other eight barcoded E-Sarbeco forward primers (incorporating the required Gibson sequence), for concat-PCR amplification. This combinatorial strategy allows to mimic the screening of 64 pseudo-candidates (**Figure 3C**). Subsequent dilutions of the synthetic SARS-CoV-2 RNA control sample were used for reverse transcription, performed with defined barcoded E-Sarbeco primers (BC33-BC36; BC38-BC40), in addition to a negative control sample devoid of viral RNA material (BC37) (dilution ID in **Figure 3C**). The real-time diagnostic assay revealed significant differences between samples presenting different viral RNA copies based on the number of aligned read counts towards the E-Sarbeco target region. Such differences, observed as early as 7 hours after DNA sequencing, revealed that samples issued from high viral RNA titers presented systematically higher amounts of aligned read counts to the E-Sarbeco target region (**Figure 3D** and **Figure S2**). Furthermore, after 4 days of DNA sequencing (72h of DNA sequencing, and 1 day more for finalizing base-calling) a total of ∼4 million reads were sequenced, corresponding to ∼38 million monomers (9.7x concatemerization factor) (**Figure 3E**). Despite this large gain on monomer sequences, only 14 thousand optimal sequences (i.e. presenting both cDNA and PCR flaking barcodes and within the expected monomer amplicon insert size) were used for the diagnostic assay, all the others presenting either too short inserts (<100nt) or missing one or both of the required barcodes, essential for the identification of the samples. Noteworthy, the analysis of ∼14 thousand optimal sequences was sufficient for discriminating samples with viral RNA titers ranging within 1 to 100 fold dilutions of the synthetic Covid-19 RNA material (**Figure 3F & 3G**); equivalent to 100 copies detected with the quantitative PCR assay (**Figure 3B)**. Indeed, the sensitivity of the assay fails to discriminate in some cases between the negative control samples (BC37) and those issued from the least viral RNA titer (1/1000 dilution; BC36) (**Figure 3F & 3G**).

The few number of optimal sequences required for performing this diagnostic assay over 64 pseudo-candidates suggests that larger candidate screenings are possible within the scale of the sequencing-depth afforded by the MinION sequencer, anticipating the upscale of this strategy to several hundred of candidates without major impact on the sensitivity.

### Real-time SARS-CoV-2 variants’ tracking over multiple pseudo-candidates

Considering the current needs on tracking SARS-CoV-2 variants’ emergence and spreading over the population, we have evolved the aforementioned real-time SARS-CoV-2 diagnostic strategy into a variants’ tracking system. Because all current described SARS-CoV-2 variants present an important number of mutations within the SPIKE gene, we have focused the real-time tracking system to this region. In fact, by concentrating the sequencing efforts towards the SPIKE gene (∼3.8kb size) we can afford to combine diagnostic and variants’ tracking into a single assay, which per se represents a major progress relative to the current strategies. In contrast to the aforementioned diagnostics effort targeting a short amplicon (E-Sarbeco), variants’ tracking over the whole SPIKE gene uses a random-hexamer sequence for the reverse transcription step, and four primers targeting the SPIKE gene at different regions (∼1kb distance among each primer) (**Figure 4A**). Like in the case of the diagnostics assay, the random-hexamer primer used for the reverse transcription present a unique molecular barcode as well as the common Gibson sequence. In contrary, the PCR primers (P1-P4) present a unique molecular barcode but they are devoid of the Gibson sequence. In fact, PCR amplification over cDNA issued from random-hexamer priming leads to amplicons of variable length (including fragments >1kb; see **Figure 4B & Figure S3**), thus the concatemerization step used on the short amplicon strategy does not appear essential. To simplify its use, PCR amplification is performed in presence of a pool of all four primers targeting the SPIKE region (P1-P4) and a Gibson primer sequence (in an equimolar concentration; i.e. 4x Gibson primer for 1x for each SPIKE targeting primer).

**Figure 4.**
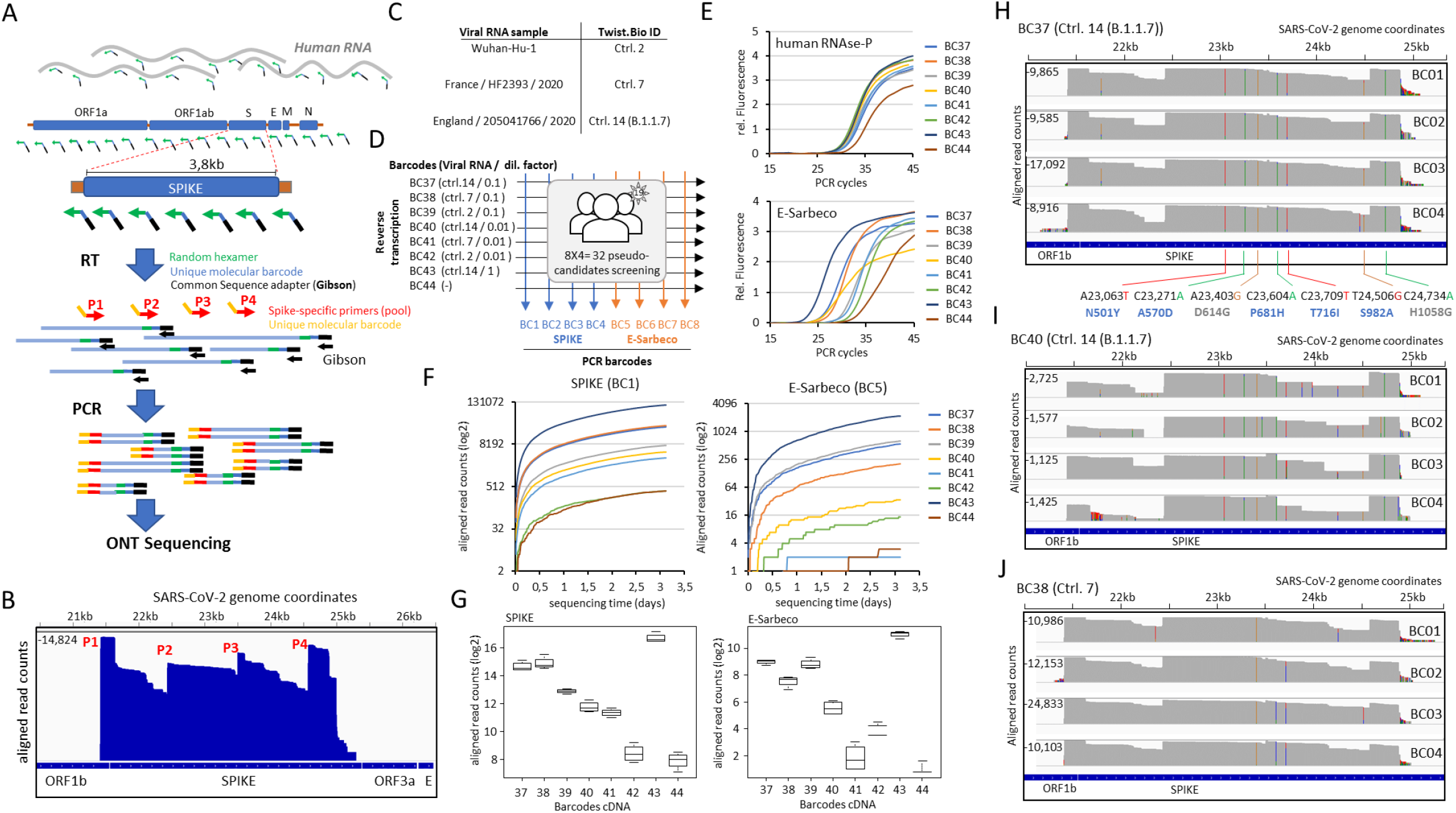
SARS-CoV-2 variants tracking within the SPIKE gene. **(A)** Scheme illustrating the SPIKE targeted strategy for variants tracking with Oxford Nanopore technology (ONT) sequencing. Reverse transcription is performed with random hexamer primers presenting unique molecular barcodes and the Gibson adapter sequence. This approach, allows to generate cDNA products of variable length covering the whole viral but also the human genome. Barcoded cDNA samples are PCR amplified in presence of four SPIKE-specific primers (each of them including a common molecular DNA barcode and spaced by ∼1kb distance) and one Gibson primer targeting the adapter sequence introduced during reverse transcription. For facilitating the assay, all four SPIKE and the Gibson primers are used within an equimolar pool. PCR amplified samples are analyzed by Oxford Nanopore MinION DNA sequencing. **(B)** Genome browser view displaying the aligned read counts obtained after ONT sequencing. The location of each of the SPIKE-targeting primers (P1-P4) is displayed and corresponds to regions presenting the highest read counts within the SPIKE gene. **(C)** Summary of the used synthetic SARS-CoV-2 viral RNA samples, corresponding to the Wuhan-Hu-1 control sample (reference strain) and two described variant strains (HF2393 and B.1.1.7). **(D)** Reverse transcription assay scheme for 7 conditions corresponding to different dilutions of viral RNA issued from the strains illustrated in (C) (BC37-BC43) and a control sample devoid of viral RNA (BC44). In all cases, 15ng of human RNA material – issued from cell lines in culture - has been also introduced. All 8 samples are pooled at the end of the reverse transcription assay and PCR amplified with either four pools of barcoded primers targeting the SPIKE gene (BC1-BC4) or four barcoded primers targeting the E-Sarbeco region (BC5-BC8), hence providing a combinatorial barcoding strategy corresponding to 32 pseudo-candidates. **(E)** Quantitative PCR targeting either the human RNAse-P gene or the viral E-Sarbeco region, performed over the reverse transcription assays described in (D). **(F)** Real-time Covid-19 diagnostic performed by RETIVAD displaying the number of aligned read counts per barcoded cDNA sample (described in (D)) for the gene SPIKE (BC1) or the E-Sarbeco region (BC5). **(G)** Boxplots illustrating the total aligned read counts associated to the gene SPIKE or the E-Sarbeco region for each of the conditions described in (D). **(H)** Variant analysis performed by RETIVAD over the barcoded sample 37 (BC37) associated to the corresponding PCR barcodes (BC1-BC4). The displayed genome browser view reveals the presence of multiple nucleotide variants (colored bars), among them seven being retrieved on all four samples (BC1-BC4) and with a significant read coverage. Five of them (A23,063T; C23,271A; C23,604A; C23,709T; T24,506G) correspond to missense mutations that were described within the B.1.1.7 UK variant strain (blue). In addition, two other missense mutations were also detected; the D614G (previously described within SARS-CoV-2 European strains) and the H1058G. **(I)** Similar to (H) but for the sample BC40, corresponding to a 10-fold lower RNA titer than that used for sample BC37. Note that the same missense mutations retrieved in (H) are observed (colored bars), and this in despite of the lower aligned read counts). **(J)** Variants detection assay over the sampled BC38, corresponding to the SARS-CoV-2 variant strain HF2393, described in France in 2020. In this case, only the missense mutation D614G is retrieved among all four samples (BC1-BC4).

To better mimic a real situation, in which the diagnostics/variants’ tracking assay is performed with samples collected from human candidates (e.g. nasopharyngeal swabs), human RNA – issued from cell lines in culture - has been combined with the synthetic viral RNA samples, such that the random-hexamer primers used for the reverse transcription assay could also provide means to count with a positive control for sample collection. Furthermore, for addressing variants tracking performance, in addition to the Wuhan-Hu-1 SARS-CoV-2 RNA control sample, we have used synthetic viral RNA samples corresponding to the highly transmissible SARS-CoV-2 variant B.1.1.7, first described in the UK in December 2020, as well as the strain HF2393 described as a particular clade circulating in France since January 2020 ^9^ (**Figure 4C**).

To perform diagnostics and variants’ tracking assay over multiple pseudo-candidates, we have used eight barcoded random-hexamer primers for reverse transcription (BC37-BC44) in presence of different viral RNA titers (consecutive dilutions) and issued from the aforementioned synthetic viral RNA strains (**Figure 4D**). To multiply the number of pseudo-candidates, we have amplified the eight barcoded cDNA products with four barcoded primers, either targeting the SPIKE gene (BC1-BC4 corresponding to pools of P1-P4) or targeting the E-Sarbeco region (BC5-BC8).

The cDNA products issued from barcoded-random-hexamer priming were first validated by quantitative PCR amplification, targeting either the human RNAse-P gene or the viral E-Sarbeco region (**Figure 4E**). Then, the PCR products generated with barcoded primers targeting SPIKE (BC1-BC4) and those targeting the E-Sarbeco region (BC5-BC8) were pooled together and analyzed by ONT sequencing. The real-time diagnostic assay for the SPIKE gene or the E-Sarbeco region, revealed significant differences between samples presenting different viral RNA copies based on the number of aligned read counts. Specifically, the negative control sample (BC44) presented systematically the least number of aligned counts for the SPIKE and the E-Sarbeco region, and in the opposite, the sample presenting the most concentrated viral RNA titers (BC43) presented the highest number of aligned reads per aforementioned target regions (**Figure 4F & 4G**).

In addition to the diagnostic outcome, RETIVAD performs a variants’ detection relative to the SARS-CoV-2 reference genome. In fact, as illustrated on **Figure 4H**, RETIVAD detected the presence of 7 significant variants within the SPIKE gene on all four samples issued from the BC37-barcoded cDNA (BC1-BC4 x BC37). Among them, five correspond to those previously described within the strain B.1.1.7 (N501Y, A570D, P681H, T716I and S982A); coherent with the fact that samples associated to the BC37 were generated with the synthetic viral RNA corresponding to such SARS-CoV-2 variant (**Figure 4D**). Similarly, variants detection performed on samples issued from the BC40-barcoded cDNA (BC1-BC4 x BC40) revealed a similar signature, in despite of their lower coverage (**Figure 4I**). In fact, samples associated to the BC40 were also generated with the synthetic viral RNA corresponding to the B.1.1.7 strain, but with a 10-fold lower titer than in the case of the BC37 (**Figure 4D**).

Finally, variants’ tracking performed on samples issued from the BC38-barcoded cDNA (BC1-BC4 x BC38; French strain HF2393) revealed a single common mutation, D614G, previously described within the French strain HF2393^9^, but also among other European strains, including the B.1.1.7, as observed within the variants detected on samples issued from BC37 and BC40 (**Figure 4H & I**).

## Discussion

Despite the current vaccination efforts, the pandemic situation anticipates the propagation of the recently described variants, as well as the emergence of new others, notably due to the major discrepancies between countries to access to optimal vaccination programs ^6^. As consequence, counting with methodologies for Covid-19 diagnostics but also variants’ tracking on a population scale remains essential.

Currently, diagnostics and variants’ tracking are performed in two steps, which per se delays the time for obtaining a complete diagnostic. Furthermore, variants’ tracking relies on full genome sequencing of SARS-CoV-2, which represent an expensive and labor-intensive effort, incompatible with population-scale assays. Finally, most of the variants’ tracking efforts, relying on the use of large and expensive sequencing instruments, requires to centralize samples processing to dedicated institutions, which are rather absent on poor countries.

To address all these pitfalls, we propose herein a methodology able to (i) perform SARS-CoV-2 diagnostic and variants’ tracking at the same time; (ii) shorten the time for accessing to the outcome of the diagnostic/variants’ detection by the use of a real-time tracking system; (iii) take advantage of the compact and affordable Oxford Nanoporer MinION DNA sequencing instruments, notably for deploying the aforementioned diagnostics and variants’ tracking platform anywhere over the world.

The proposed variants’ tracking focus on the analysis of the SPIKE gene instead of the whole SARS-CoV-2 genome. In fact, the SPIKE gene has shown to concentrate the majority of the currently discovered variants, which can be rationalized by the fact that it codes for the protein required for the interaction with the human cells. Noteworthy, most of the current vaccines against the Covid-19 are also based on the SPIKE region. Hence, concentrating variants’ tracking towards this gene appears an optimal compromise to increase the number of candidates to screen at once. This philosophy is shared by a recent work describing the HiSpike method, performing variants’ detection within the SPIKE gene with the help of the small Illumina MiSeq instrument ^10^. Illumina instruments being a short fragments sequencing approach, HiSpike requires 42 primer pairs generating amplicons of 400 nucleotides of length for covering the SPIKE region. In contrary, our proposed strategy, based on the use of long DNA fragments, compatible with the Nanopore DNA sequencing technology, relies on 4 primers targeting the SPIKE region, one random primer for the cDNA step and a common Gibson sequence. In fact, a strategy for SARS-CoV-2 diagnostic and variants’ surveillance, dedicated to be deployed anywhere on the world does not only requires to count with low cost instruments, but also involve a reduced number of reagents for allowing its use on a long-standing manner.

For all these reasons, we are convinced that our proposed strategy, accompanied by the dedicated computational solution RETIVAD, can represent a game-changer approach for tracing variants’ progression in the following months.

## Data Availability

all data are available under request.

## Acknowledgements

We gratefully acknowledge Corinne Cruaud and her team for the technical training on Oxford Nanopore sequencing. We also thank Veronique de Berardinis and Patrick Wincker for the logistics support required for this project.

## Author contributions

Conceptualization: M.A.M.P. Methodology: J.L.P. and M.A.M.P. Software development: F.S. and S.E. Scientific evaluation: M.A.M.P. Writing, review, and editing: J.L.P., F.S., and M.A.M.P. Funding acquisition: M.A.M.P.

## Additional Information

Correspondence and requests for materials should be addressed to M.A.M.P.

## Methods

### SARS-CoV-2 synthetic RNA

The following Synthetic SARS-CoV-2 RNA control samples, delivered at a concentration of 1 million copies per microliter has been used in this study:

- Wuhan-Hu-1 (GeneBankID: MN908947.3); Twist Biosc. ID: Ctrl2.
- France/HF2393/2020 (EPI_ISL_418227); Twist Biosc. ID: Ctrl7.
- England/205041766/2020 (EPI_ISL_710528); Twist Biosc. ID: Ctrl14 (B.1.1.7).

### Barcode primer design

Primers used for reverse transcription are composed by (i) a fragment of the Gibson sequenced (redGibs: GAAGAACCTGTAGATAACTCGCTGT), (ii) a barcode sequence issued from the ONT PCR Barcoding Expansion 1-96 kit (EXP-PBC096) and (iii) the target sequence. For targeting the E-Sarbeco region within the SARS-CoV-2 genome, the reverse primer sequence provided by Corman et al ^8^, has been used:

E_Sarbeco_R: ATATTGCAGCAGTACGCACACA

For SPIKE-centered variants tracing, the target sequence has been replaced by a random hexamer, notably to decrease the number of required primers during the assay.

The following barcoded primers targeting the E-Sarbeco region were used for reverse transcription:

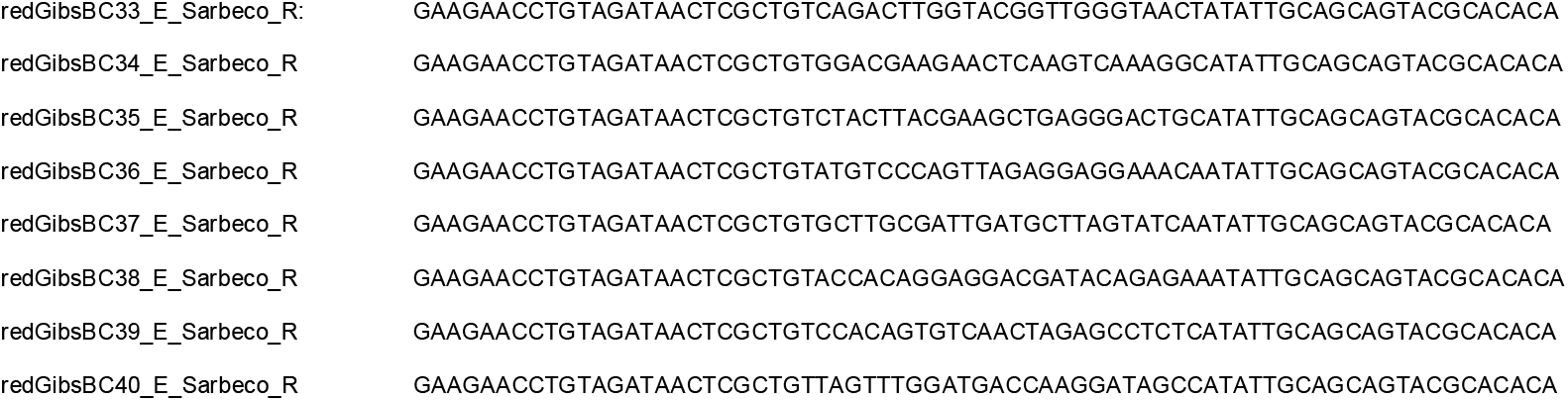

The following barcoded random-hexamer primers were used for reverse transcription:

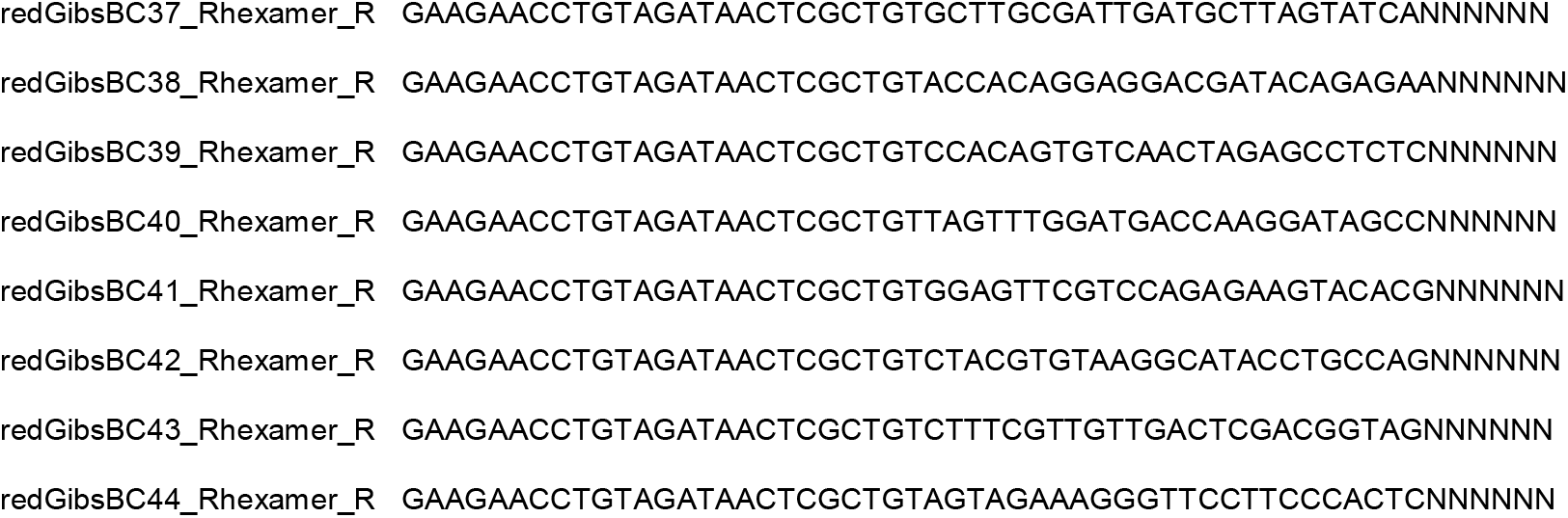

For PCR amplification a second set of barcoded primers were designed presenting the following structure: (i) A reverse-complementary Gibson fragment (revGibs: ACAGCGAGTTATCTACAGGTTCTTCAATGT), (ii) a barcode sequence issued from the ONT PCR Barcoding Expansion 1-96 kit (EXP-PBC096), and (iii) the target sequence. For targeting the E-Sarbeco region within the SARS-CoV-2 genome, the forward primer sequence provided by Corman et al ^8^, has been used:

E_Sarbeco_F:ACAGGTACGTTAATAGTTAATAGCGT

The following barcoded primers targeting the E-Sarbeco region were used in this study:

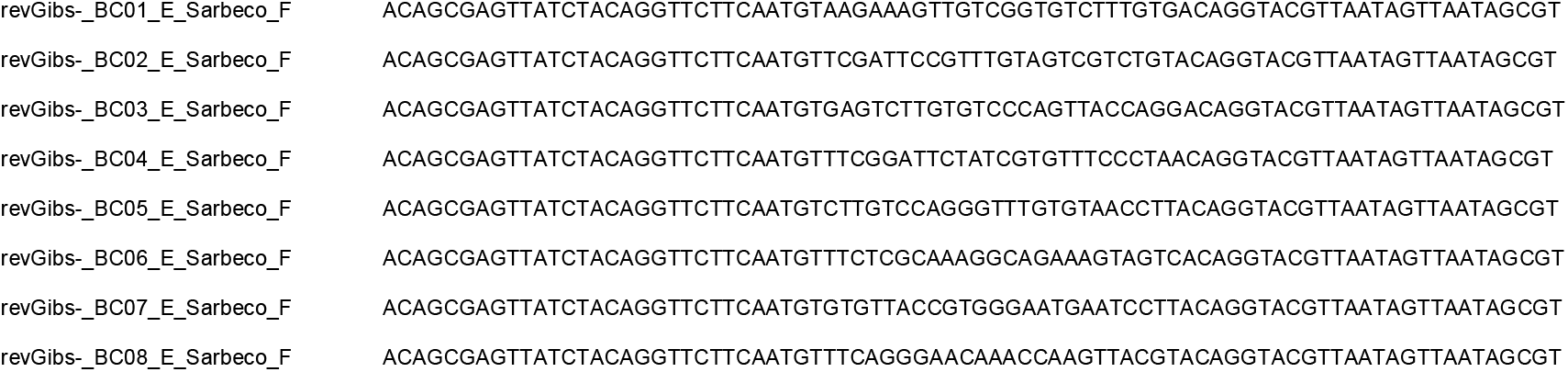

For SPIKE-centered variants tracing, the reverse-complementary Gibson sequence has been excluded, notably because the PCR amplification performed over random-hexamer generated cDNA templates generate large fragments (>1kb). In contrary, the amplicon product issued from E-Sarbeco targeting oligonucleotides gives rise to short amplicons – compatible with quantitative PCR assays -, thus requiring concatemerization prior nanopore sequencing.

With the aim of covering the SPIKE gene (∼3.8kb), the following four primers targeting the SPIKE region with ∼1kb interval within them were designed:

Spike1_F: AGGGGTACTGCTGTTATGTCT

Spike2_F: TGCACTTGACCCTCTCTCAG

Spike3_F: GCAGGCTGTTTAATAGGGGC

Spike4_F: TGCAGACATATGTGACTCAACA

The following Spike targeting primers including the corresponding barcode sequences were used for this study:

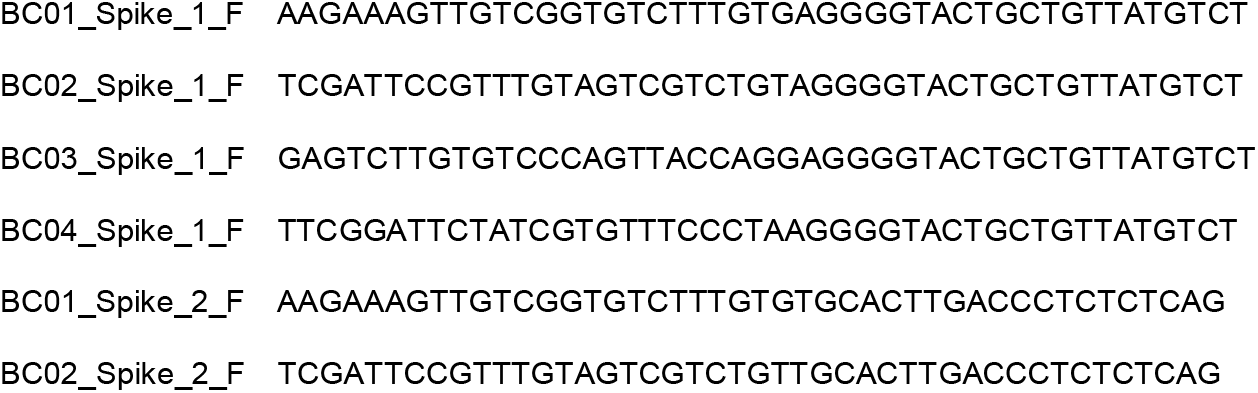
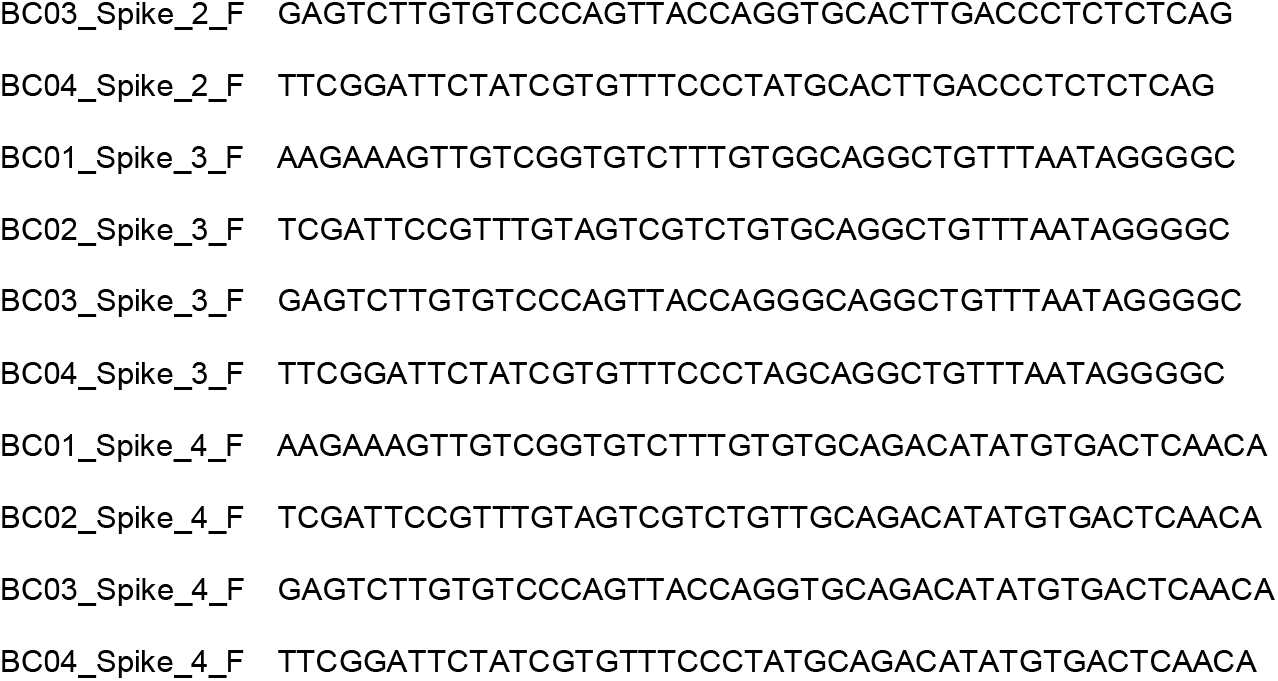

### Reverse transcription

Reverse transcription assays were performed using SuperScript^™^ IV Reverse Transcriptase (Thermofisher Scientific; 18090200; 0.5 ul superscript IV (200U/ul)) with a standard reaction volume of 20ul including 0.5ul of super RNAseIn (Thermofisher Scientific; AM2694; 20U/ul), 1ul DTT (0.1 M), 4 ul SSIV buffer (5X), 0.5 ul dNTP (10 mM each), 1 ul of synthetic SARS-CoV-2 RNA, 1 ul of 2uM gene-specific-barcoded oligonucleotide (e.g. E-Sarbeco barcodes bc33-bc48; 0.1uM final concentration) or 1 ul of 50uM random hexamer-barcoded oligonucleotide (final concentration: 2.5uM). For assays including random hexamer-barcoded primers, 1ul of human RNA (15 ng/ul; collected from cell lines in culture) were added to the assay together to the synthetic SARS-CoV-2 RNA and prior adding the random hexamers. Synthetic SARS-CoV-2 RNA was added at a copy number of 1 million (dilution factor, d.f: 1), 100 thousand (d.f: 0.1), 10 thousand (d.f: 0.01) and 1 thousand (d.f: 0.001) per reaction respectively. Note that these consecutive dilutions give rise after downstream dilutions to the number of copies of 10 thousand, 1 thousand, 100 and 10 respectively, analyzed by quantitative PCR (**Figure 3B**).

Multiple reverse transcription assays – each of them performed in presence of a defined barcoded primer - were incubated at 50°C during 30 minutes, then at 80°C 10 minutes. Barcoded cDNA samples were pooled (8 samples at once) and cleaned with SPRIselect reagent (Beckman; B23318) with a ratio of 0.5x. Cleaned cDNA is recovered on 80ul (half of the total initial cleaned volume; i.e. 2x concentration). 40ul of this pooled & cleaned cDNA will be used for PCR amplification (5ul x 8 PCR reactions; see below), while the remaining material is used for quantitative PCR evaluation.

### SARS-CoV-2 Quantitative PCR validation

Barcoded-cDNA material has been validated by quantitative PCR assay with the following primer sequences:

Primers targeting region on E gene of SARS-Cov-2 (annealing temp: 58°C:

**E_Sarbeco_F:** ACAGGTACGTTAATAGTTAATAGCGT

**E_Sarbeco_R:** ATATTGCAGCAGTACGCACACA

Primers targeting the human gene RnaseP (annealing temp: 60°C):

**Human_RNASE_P-F:** AGATTTGGACCTGCGAGCG

**Human_RNASE_P-R:** GAGCGGCTGTCTCCACAAGT

Quantitative PCR assays were performed with QuantiTect SYBR Green PCR Kit (ref: 204145) over a 1/10 diluted cDNA material.

### Concat PCR amplification and Nanopore sequencing

PCR amplification targeting the E-Sarbeco amplicon is performed on 25ul final volume including 12.5ul of Phusion Hot Start II High-Fidelity PCR Master Mix (ThermoFisher ref: F565), 2.5ul of 1uM barcoded-E-Sarbeco targeting primer (BC1-8), 2.5ul of 1uM short Gibson primer (AGAACCTGTAGATAACTCGCTGT) and 5ul of the pooled & cleaned cDNA. For short-amplicon E-Sarbeco PCR amplification the following thermal cycling program is used:

1. 98°C 30 seconds
2. 98°C 10 seconds
3. 61°C 20 seconds
4. 72°C 30 seconds
5. Repeat 2-4 steps 25x
6. 98°C 10 seconds
7. 61°C 20 seconds
8. 72°C 2 minutes
9. Repeat 6-8 steps 20x
10. 72°C 10 minutes
11. Keep at 12°C

For PCR amplification targeting SPIKE and E-Sarbeco on random-hexamers generated cDNA material the assay is performed with a pool of barcoded SPIKE primers and shortGibson. For 100ul mix 10ul of barcoded primer Spike1 (10uM), 10ul of Spike2 (10uM), 10ul of Spike (10uM), 10ul of Spike4 (10uM) and 40ul of ShortGibson (10uM) completed with distilled water is prepared. Then the PCR amplification is performed on 25ul final volume including 12.5ul of Phusion Hot Start II High-Fidelity PCR Master Mix (ThermoFisher ref: F565), 1.25ul of pooled barcoded-SPIKE & shortGibson primers (0.05uM final concentration per SPIKE targeting primers and 0.2uM final concentration for ShortGibson) and 5ul of pooled & cleaned cDNA.

For short-amplicon E-Sarbeco PCR amplification the following thermal cycling program is used:

1. 98°C 30 seconds
2. 98°C 10 seconds
3. 61°C 45 seconds
4. 72°C 2 minutes
5. Repeat 2-4 steps 39x
6. 72°C 10 minutes
7. Keep at 12°C

In both cases (E-Sarbeco Concat-PCR or SPIKE PCR amplification), PCR products were verified by TapeStation automated electrophoresis (Genomic DNA ScreenTape; Agilent ref: 5067-5365).

Multiple PCR assays are performed in presence of different barcoded primers, such that the combination with the barcoded cDNA material allows to generate a large complexity. Specifically, for E-Sarbeco targeting assay, 8 barcoded contact-PCR assays are pooled (8×25=200ul) and cleaned with SPRIselect reagent (Beckman; B23318) with a ratio of 0.5x. Similarly, for SPIKE and E-Sarbeco PCR assays issued from random hexamer cDNA material, 4 barcoded PCR assays for each target regions (SPIKE: BC1-4; E-Sarbeco: BC5-8) were pooled (4×2×25=200ul) and cleaned with SPRIselect reagent (Beckman; B23318) with a ratio of 0.5x.

Pooled & cleaned material is recovered on 50ul and used for Nanopore sequencing library preparation (Nanopore Ligation Sequencing Kit: SQK-LSK109). DNA libraries were sequenced on Nanopore FLO-MIN106D flowcells (R9) with the MinION Mk1C instrument (72h sequencing; high-accuracy base calling).

### Real-time SARS-CoV-2 diagnostics and variants tracking

Real-time processing of the fastq files generated by the Mk1C instrument during sequencing is performed with our in-house developed RETIVAD (Real-Time Variants Detector) software. RETIVAD collects each ten minutes the fastq files generated during sequencing from the Mk1C instrument and (i) search for Gibson sequences within the sequenced reads (regex query (https://pypi.org/project/regex) with maximum 4 mismatches accepted by default); (ii) split sequenced reads on monomers – delimited by the presence of the identified Gibson sequences; (iii) search for barcode sequences introduced during the reverse transcription (cDNA barcodes: BC33-BC48; regex query); (iv) search for barcodes introduced during the PCR amplification (PCR barcodes: BC1-BC8; regex query); (v) collect the sequences between the cDNA and PCR-associated barcodes for their alignment towards the SARS-CoV-2 reference genome (Wuhan-Hu-1 genome: NC_045512; BWA aligner following nanopore parameters). Considering that steps (iii) to (v) are performed within the retrieved monomers in step (ii), RETIVAD matches cDNA/PCR barcode combinations to aligned outcomes, thus providing a diagnostic outcome for the associated pseudo-candidate. Due to the low number of sequences retrieved at intervals of 10 minutes, and the relative short size of the SARS-CoV-2 genome (∼29kb), RETIVAD is able to provide such diagnostic outcome within such 10 minutes interval. Hence, at the next round of data collection, RETIVAD reiterates on the aforementioned steps over the newly collected fastq files and appends the outcome to the previous results. The continuous gain on aligned read-counts per region of interest (e.g. E-Sarbeco amplicon sequence, or SPIKE gene; defined as entry parameters) is visualized within scatterplot view (png file) updated during the processing (RETIVAD produces a png file per detected PCR barcode monomers; thus allowing to generate scatterplots displaying as many lines as detected cDNA barcode monomers; **Figure S4**).

In addition to the diagnostic outcome, RETIVAD has been designed for performing real-time variants detection. For it, the <u>racon package</u> (version 1.4.20) is used for raw de novo DNA assembly, followed by the alignment with <u>Minimap2</u> (version 2.17). <u>Medaka</u> (version 1.2.5) is used to create a consensus sequence and variant calls from the aligned data (> 20 aligned reads required as default for variants calling). RETIVAD generates BAM files per cDNA/PCR barcode combination harboring base conversion information and their associated coverage (for visualization with Genome browsers like <u>IGV</u>). Furthermore, summary files per cDNA/PCR barcode combination are also generated, providing the genomic position in which the base conversion has been detected, the identity of the converted bases as well as their associated coverage. Like in the case of the diagnostics processing, variants detection is performed in real time, thus providing the possibility to the user to visualize the results and decide whether sequencing requires still to be performed or whether the collected information is sufficient to conclude the diagnostics/variants tracing for the corresponding candidates.

RETIVAD generates a final summary report for the diagnostics outcome when no new fastq files are available after 40 minutes from the last data collection and processing.

### Data and Software availability

All genome sequences included in this study are available under request. RETIVAD is freely available at https://github.com/SysFate/retivad

