## Supplementary Figures for "Real-time SARS-CoV-2 diagnostic and variants tracking over multiple candidates using nanopore DNA sequencing"

### Supplementary Material

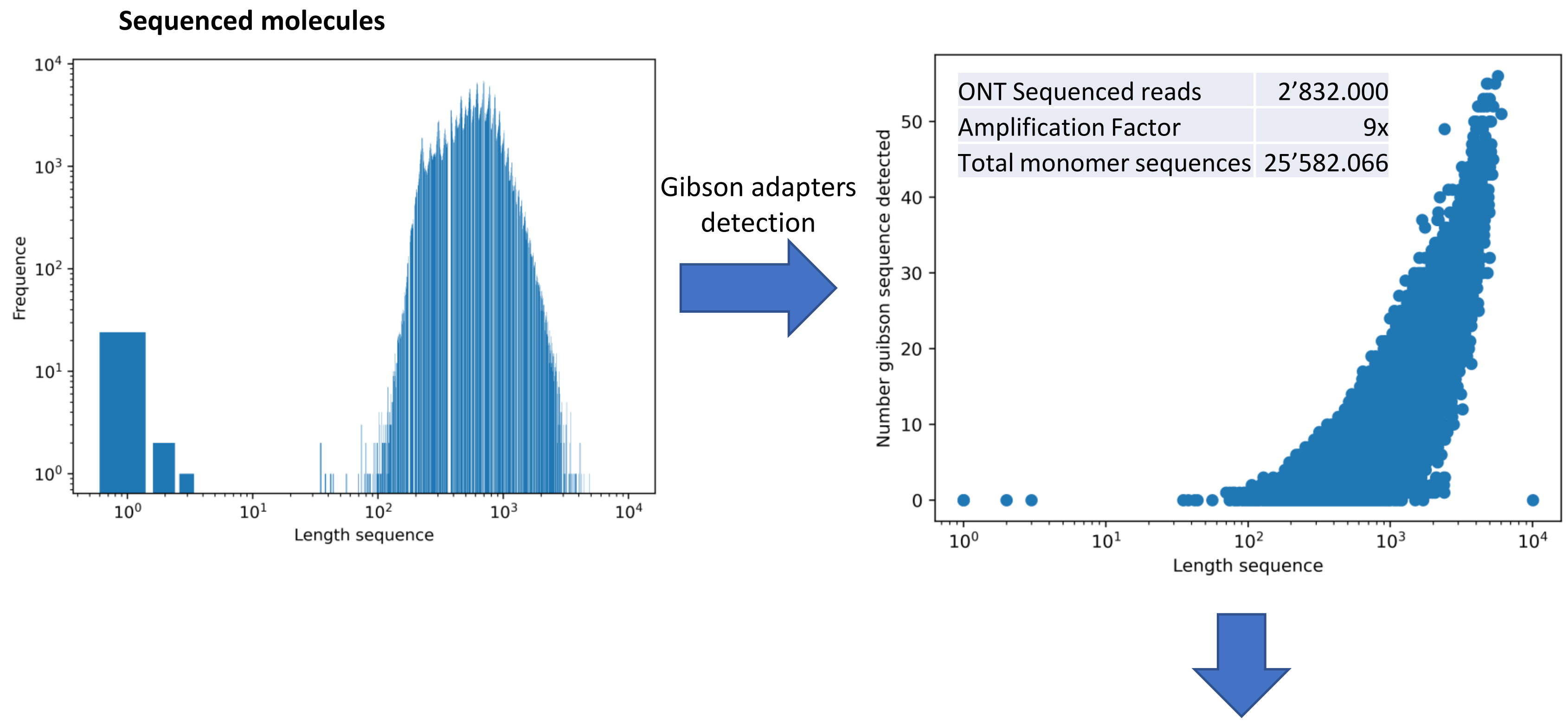


**Figure S1. monomer detection within Nanopore sequenced reads issued from concat PCR.**
***Top-left:*** Distribution frequency of the length of the sequenced molecules. ***Top-right:*** Scatterplot illustrating the length of the sequenced molecules and the number of Gibson sequences retrieved by RETIVAD. Inset: Total number of sequenced reads, total number of monomer sequences inferred after Gibson adapters detection. The amplification factor corresponds to the ratio between the number of monomer sequences and the initial ONT sequenced reads. ***Bottom-center***: Distribution frequency of the length of the sequences between Gibson adapters. Captured sequenced within Gibson adapters are further classified on those presenting a “short length” (defined by the expected size of the amplicon; i.e. 125nt for Esarbeco) and those presenting in addition both cDNA and PCR barcodes ( a & b respectively). Alignment versus SARS-CoV-2 reference genome is only performed over the last group of sequences.

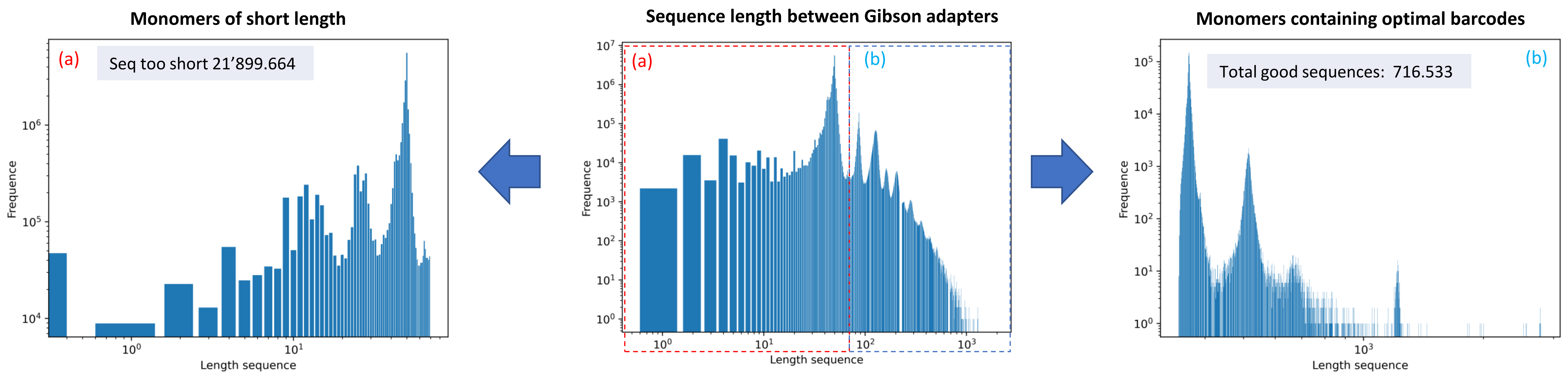

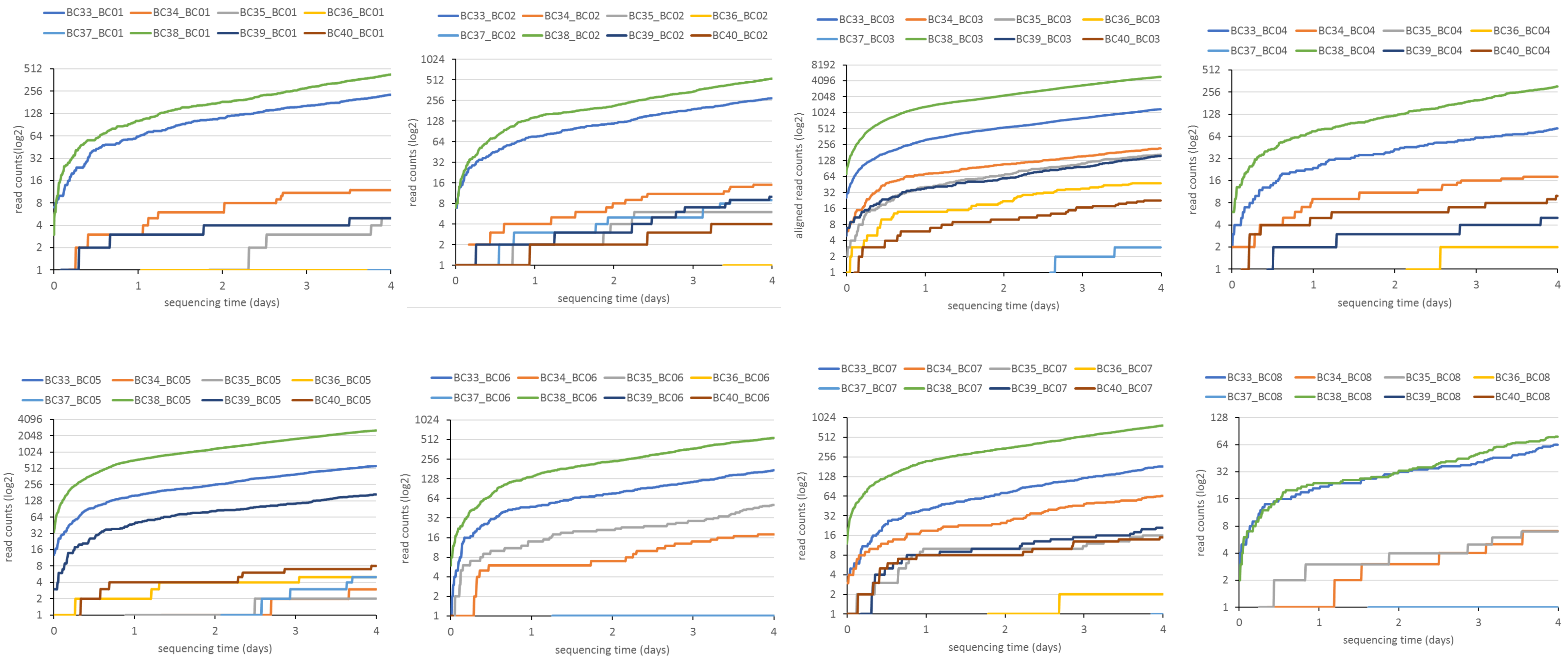


**Figure S2. Real-time display illustrating the number of aligned read counts associated to the E-Sarbeco region per dual barcode combination assessed during Nanopore sequencing.**

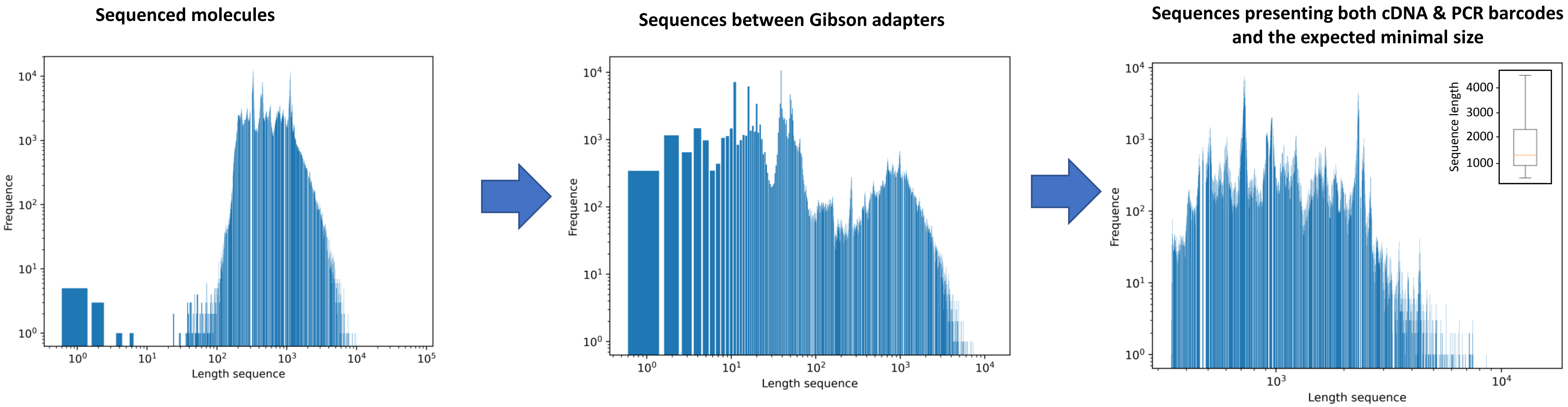


**Figure S3. monomer detection within Nanopore sequenced reads issued from random hexamer-based reverse transcription and PCR amplification**.
***Left panel:*** Distribution frequency of the length of the sequenced molecules. ***Middle panel:*** Distribution frequency of  the length of the sequences between Gibson adapters. Right panel: Same as the middle panel, but concerning molecules presenting in addition both the cDNA & PCR barcode sequences. Note that in contrast to the molecules observed for the short amplicon concatemerization assay (Figure S1), these optimal inserts present lenghts >  1kb (see boxplot inset).

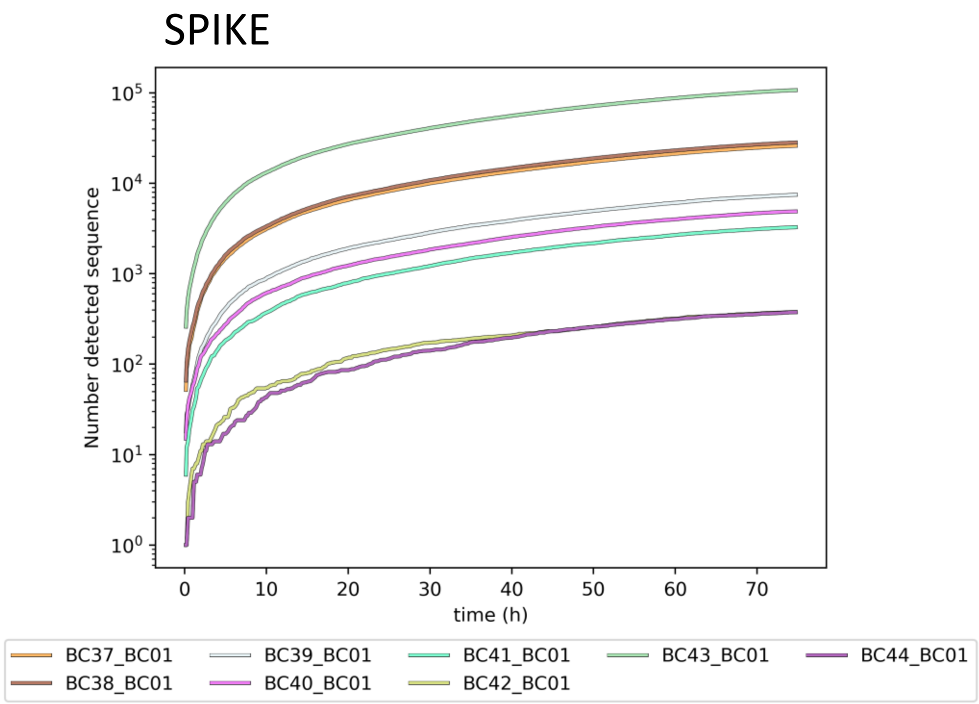


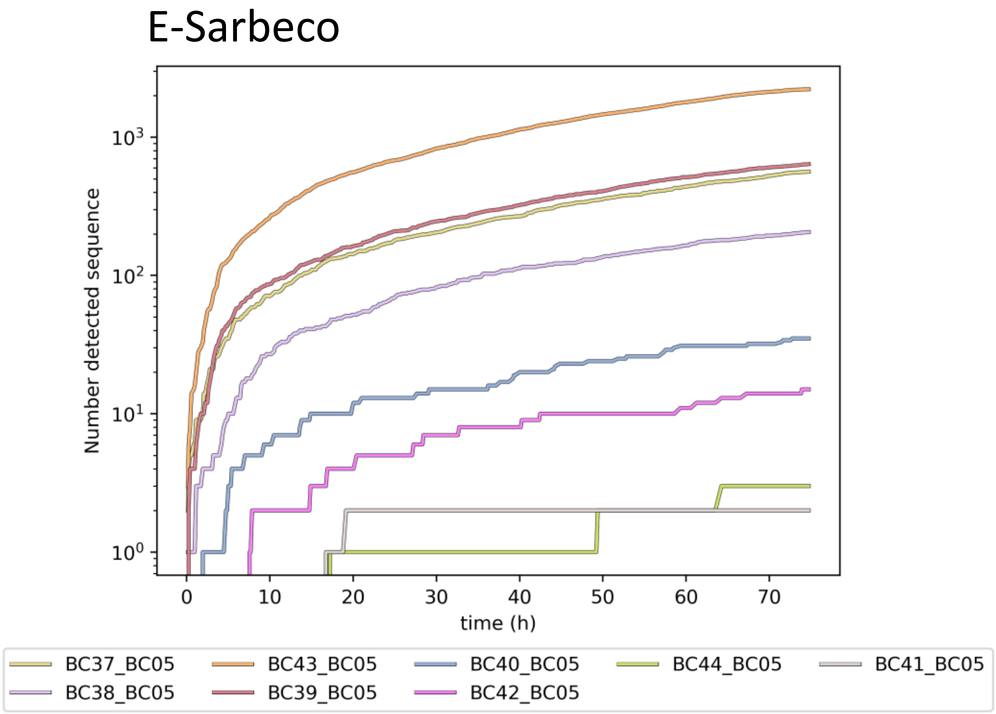


**Figure S4. Real-time diagnostics tracing generated by RETIVAD.** Representative images (png format) generated by RETIVAD during nanopore sequencing of libraries issued from PCR amplification assays targeting the SPIKE gene as well as the E-Sarbeco viral region (Figure 4).
